# Hospitalisation and healthcare burden of respiratory syncytial virus in adults over 50 years in France

**DOI:** 10.64898/2026.01.13.26344008

**Authors:** Mathias Vacheret, Jessica Y. El Khoury, Stéphane Fiévez, Emmanuelle Blanc, Laurence Watier, Jean-Sebastien Casalegno, Hervé Lilliu, Philippe Vanhems, Louise Baschet, Clélia Bignon-Favary, Pippa McKelvie-Sebileau, Lauren Inchboard, Paul Loubet

## Abstract

**Background and objective:** Respiratory Syncytial Virus (RSV) can cause severe illness in adults, leading to respiratory and non-respiratory complications, functional decline, hospitalisation, and death. This study describes French patients aged ≥50 years hospitalised with RSV (2015–2022) and their care pathways, including hospitalisation and outpatient healthcare use and costs.

**Methods:** Data were extracted from the French National Health Data System (SNDS). Patients were classified into four risk groups (high: mmunocompromised; medium: underlying predisposition; other; no comorbidities) and four age groups (50–59, 60–64, 65–74, ≥75 years). Healthcare use within 30 days post-hospitalisation (laboratory tests, imaging, pharmacy, GP visits, rehospitalisations) and costs were analysed across four periods: 60–30 days pre-hospitalisation, 30 days pre-hospitalisation, index hospitalisation, and 30 days post-hospitalisation.

**Results:** We identified 15,509 RSV-related hospitalisations for 15,169 adults ≥50 years, mainly aged 65–74 (20.9%) or ≥75 years (61.7%). Median age was 80 years; 15.9% were high risk, 71.4% medium risk, 4.8% other, and 7.8% had no comorbidities. Intensive care was required in 25.4% of cases. In-hospital mortality was 8.5%, with an additional 3.8% dying within 30 days post-discharge. Rehospitalisation occurred in 17.8% of patients, nearly half for cardiorespiratory causes. Mean index hospitalisation cost was €6,876 (SD €9,346), and post-hospitalisation costs increased across all ages and risk profiles compared to the control period.

**Conclusion:** RSV imposes a substantial hospitalisation and cost burden in adults ≥50 years, especially older patients and those with comorbidities. Expanded preventive vaccination strategies could help reduce this impact.

## Background

Long recognised as a major paediatric pathogenic infection (1), respiratory syncytial virus (RSV) is also an important and underestimated health risk in older and high-risk adults (2–4). RSV infection presents a wide clinical spectrum, from asymptomatic infection and mild respiratory illness, to severe pneumonia or acute respiratory distress, and it exacerbates underlying chronic respiratory and cardiovascular conditions including acute decompensated heart failure (5–7). In older populations, particularly those with chronic diseases, RSV infection substantially increases healthcare resource utilisation, hospitalisation rates, cardiovascular complications, functional impairment, and mortality (3,5–9). Beyond the acute hospitalisation phase, RSV can lead to sustained functional decline, increased dependency, institutionalisation, and elevated rehospitalisation risk among older adults (8,10). However, such longer term outcomes remain inadequately quantified in the current literature.

The hospital burden of RSV among adults in France has been estimated through French hospital discharge data (Programme de Médicalisation des Systèmes d’information, PMSI) over 2012 and 2021, identifying 12,987 adult RSV-related hospitalisations, with a mean age of 74.1 ± 16.4 years and 57% of patients over 75 years old (11). RSV-attributable hospitalisation rates over 2010-2020 for adults ≥ 65 years in France have been recently reported at 174 per 100,000 person-years for respiratory causes, and 202 for narrowly-defined cardiorespiratory causes (12). However, substantial variability is noted. While one recent study estimated adjusted rates for adults over 75 years at 85 to 221 per 100,000 (13), another study using extrapolation methods estimated the incidence of RSV-associated hospitalisations among adults aged 60 years and older at 289 per 100,000 (14). In addition to variability, reported RSV hospitalisation incidences are likely to be underestimated due to inconsistent testing practices, symptoms similar to those caused by other respiratory viruses (15–17) and incomplete RSV coding and underreporting (11). Further, studies examining community-level epidemiological impacts, ambulatory care utilisation, and comprehensive economic implications beyond paediatric populations remain limited (18–20). European prospective cohort studies recently reported average healthcare costs per RSV episode ranging from €30.8 to €78.1 per patient, but these estimates were limited to non-hospitalised cases and did not include France (10,21). Mean hospital costs for RSV-related respiratory episodes in France are reported at €5,199 for adults aged 50+ (12), and the average cost per RSV-coded hospitalization in adults aged 75 and above varied from €5,021 to €5,706, depending on the year (22). However, to date, no study has comprehensively measured the economic impact including direct and indirect costs of RSV infection in adults both during and after hospitalisation.

## Methods

Our study aims to describe the economic impact through healthcare claims, and overall burden of RSV infections among French adults aged ≥50 years hospitalised with RSV from 2015 to 2022.

Study design and data sources: Our retrospective study is a claims database analysis of healthcare data from the French National Health Database (SNDS). The SNDS contains hospitalisation information from the PMSI database for all public and private hospitals as well as healthcare use data from national claims database (*Datamart de Consommation InterRegimes* - DCIR). It is one of the largest and most exhaustive in Europe, with ten years of follow-up for around 99% of the French population (23).

Participant characteristics: The target population consisted of all adult patients over 50 years of age who were hospitalised at least once with RSV between 1^st^ June 2015 and 31^st^ July 2022. This period includes seven epidemiological years, defined as July to the following June according to a previous epidemiology study (24), from 2015-2016 to 2021-2022. Hospitalisation with RSV infection was defined as a hospital admission (with or without overnight stay) in Medical Surgery and Obstetrics (MSO) with any of the following four ICD10 codes as main, related or associated diagnostic codes: B97.4 (RSV as the cause of disease classified in other chapters); J12.1 (pneumonia due to RSV); J20.5 (acute bronchitis due to RSV); or J21.0 (acute bronchiolitis due to RSV)(11). Any rehospitalisations with RSV occurring in MSO or follow-up and rehabilitation care (*Soins de Suite et Readaptation*/ *Soins Médicaux et de Réadaptation)* (SSR/SMR) within 30 days of the initial hospitalisation discharge were considered the same RSV hospitalisation episode. In these cases, the time between discharge and readmission was not included in the duration of the index hospitalisation. Rehospitalisations after 30 days were considered new hospitalisations (even when they were for the same patient). The index date corresponds to the first admittance in the MSO and discharge date as the final discharge from MSO or SSR per hospitalisation episode.

Procedure and statistical analysis: Hospitalisation burdens according to patient age and risk subgroups were described. Four age categories/strata were defined: 50-59 years, 60-64 years, 65-74 years and 75 years or over. Results for the total population were also reported. To detect preexisting comorbidities, we applied algorithms taking into account long-term disease status and healthcare use (inpatient and outpatient claims) over a five-year historic period prior to the index hospitalisation date. The evidence of comorbidities over this five-year period was used to classify patients into four risk groups, from most to least severe, defined in consultation with a group of experts based on a transposition of populations at risk for influenza and pneumococcal infections: 1) High-risk patients corresponding to immunocompromised patients with at least one comorbidity from the following: asplenia or hyposplenia, hereditary immune deficits, HIV infection, chemotherapy-treated cancer, solid organ transplant, hematopoietic stem cell transplant, chronic autoimmune or inflammatory diseases treated with immunosuppressive or biologic drugs, or nephrotic syndrome; 2) Medium-risk, corresponding to non-immunocompromised patients with underlying predispositions including chronic respiratory disease (COPD, emphysema, severe asthma under continuous treatment), cyanotic heart disease, heart failure, renal disease, chronic liver disease, diabetes, obesity or multiple sclerosis; 3) other generalised risk factors, such as sleep apnoea, tobacco use, alcohol use, malnutrition, Alzheimer’s disease, Parkinson’s disease or dementia; and 4) patients without evidence of former pathologies. Risk groups were mutually exclusive and if a patient had more than one condition, the most severe risk group classification was retained. Algorithms for each pathology are presented in Table S1.

Index hospitalisations were described according to month of admission, geographic area, admission and discharge sources, length, and whether they included admission to follow-up and rehabilitation care (SSR/SMR) or in the intensive care unit (ICU), including continuous care, intensive care and/or resuscitation.

The date of death was recorded in the DCIR and, if it occurred in a hospital, in the PMSI. If the dates differed, the date in the DCIR was considered to take precedence. Mortality, admission to a nursing home and rehospitalisation rates at 30 and 90 days after the index hospitalisation were extracted for patients alive at discharge.

For patients alive at discharge, healthcare use was evaluated over three periods either side of the index hospitalisation: a pre-infection reference period from 60-30 days before the index hospitalisation date; a pre-hospitalisation period from 29 days before the index date to the index date; and a post-hospitalisation period from the discharge date (included) to 30 days after, as a commonly used metric (25). The following healthcare use was described: GP and specialist visits (community care), chest X-rays and laboratory tests (from community care or hospital) and respiratory drugs, painkillers, antiviral medication and antibiotics (from community care). Global healthcare costs were calculated over each period according to the national health insurance perspective, and outpatient and inpatient costs were distinguished to identify what type of healthcare that drive the highest costs. A linear mixed effects model was estimated to the cost difference between the post-hospitalisation period and the reference period, according to the epidemiological year and patient characteristics, sex, age category and risk group. Residual variance was estimated for each risk group to account for greater variance in the high-risk group (immunocompromised patients). A random effect of the geographical area of index hospitalisation was included and interactions between age and risk groups were tested. Marginal means were reported for each modality, and summary statistics of the model are provided in the supplementary materials.

## Results

### Hospitalisations with respiratory syncytial virus infections between June 2015 and July 2022

From 2015-2022, a total of 15,509 hospitalisations with RSV were observed for 15,169 adults over 50 years of age in France (Table 1), among which, 233 did not include an overnight stay. They occurred primarily in January (33.6%) or December (24.2%) (Figure 1, Table S2 and Table S3, supplementary materials). The number of hospitalisations varied annually, between 603 and 4,252 per epidemiological year (Table S4, supplementary materials) and according to geographical area (Tables S2 and S3). The areas with the greatest number of hospitalisations were “Ile-de-France” (Paris and surrounding area) and the “Auvergne Rhone Alpes” area (Figure S1, supplementary materials).

**Figure 1.**
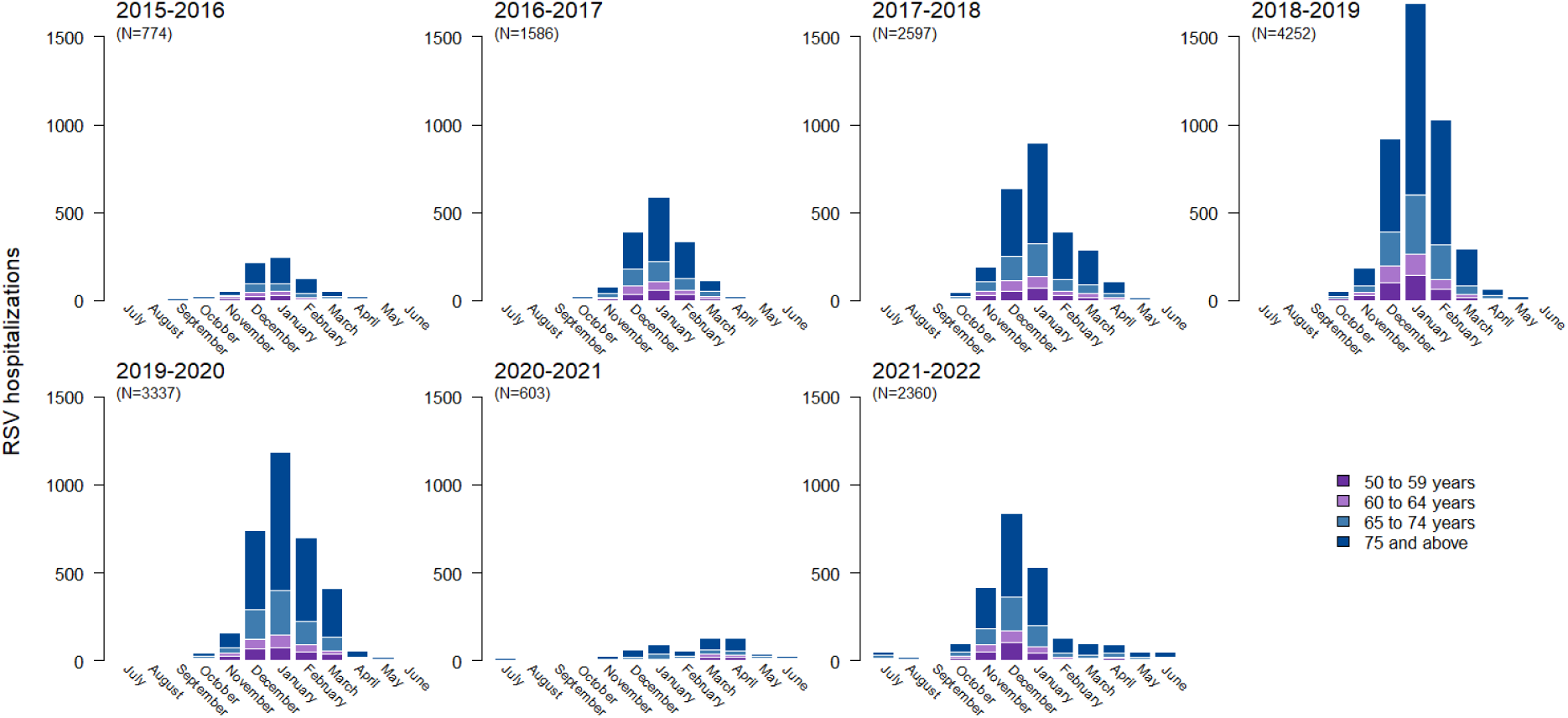
Hospitalisations for respiratory syncytial virus by month for each epidemiological year, by age group.

**Table 1.**
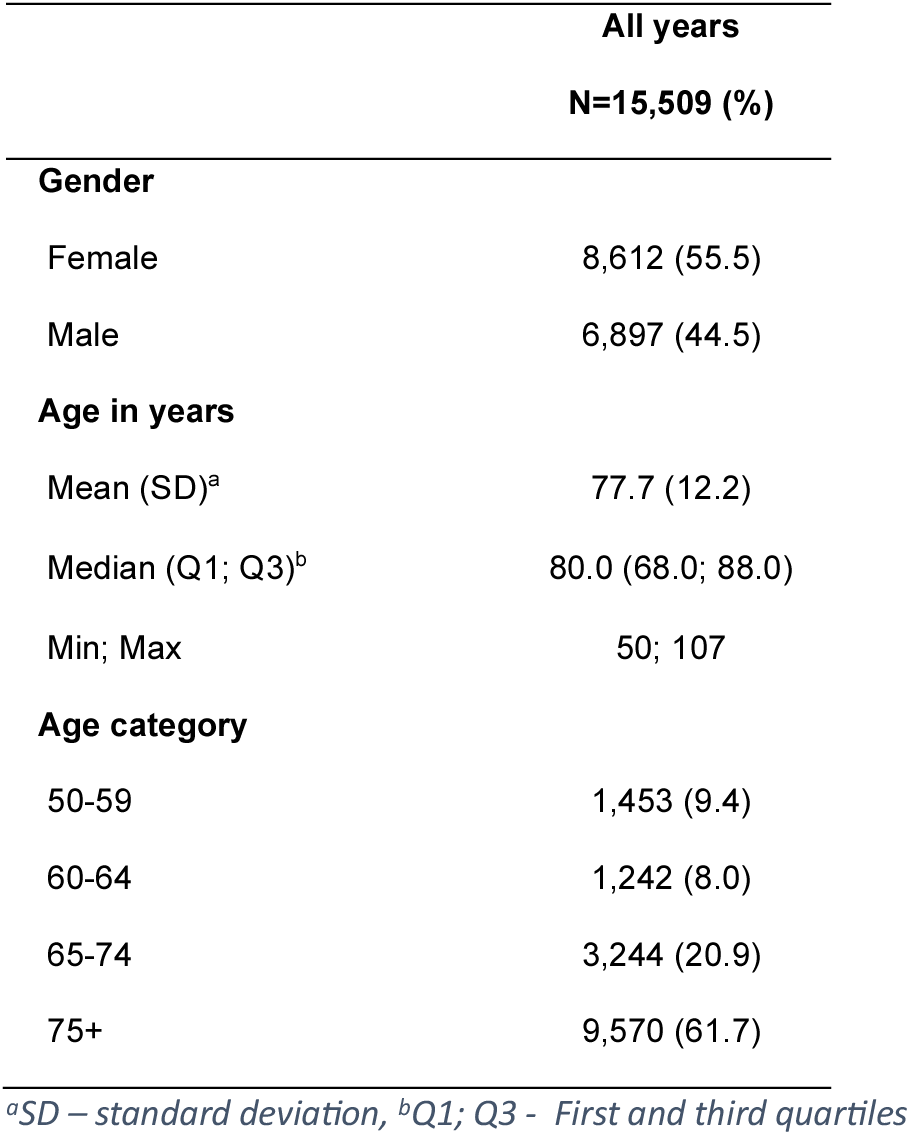
Characteristics of adult patients over 50 years hospitalised for respiratory syncytial virus in France between 2015 and 2022.

### Description of the hospitalised population

Just over half of the patients hospitalised with RSV were women (55.5%), the median age was 80.0 years, 20.9% were 65 to 74 years and 61.7% were 75 years or older (Table 1). Most patients had at least one comorbidity (92.2%). There were 15.9% high-risk immunocompromised patients (primarily receiving chemotherapy for cancer, 47.5%) and 71.4% medium-risk patients with underlying predisposing comorbidities (mostly chronic respiratory disease (55.8%), heart failure (52.4%), diabetes (33.8%) or chronic coronary disease (22.1%)). The proportion of high-risk patients was greatest among patients aged 50-59 years (32.3%) and the proportion of medium-risk patients was greatest among patients aged 75 years or older (79.0%) (Table 2). Only 7.8% of hospitalised patients had no evidence of any comorbidities. On average, patients had 2.6 (SD 1.7) comorbidities.

**Table 2.** Comorbidities of patients hospitalised for respiratory syncytial virus in France between 2015 and 2022 by age group.

|  | 50 to 59 years<br>N=1,453 (%) | 60 to 64 years<br>N=1,242 (%) | 65 to 74 years<br>N=3,244 (%) | 75 years or over<br>N=9,570 (%) | All patients (50y+)<br>N=15,509 (%) |
| --- | --- | --- | --- | --- | --- |
| **No comorbidities** | 152 (10.5) | 99 (8.0) | 244 (7.5) | 718 (7.5) | 1,213 (7.8) |
| **Immunocompromised patients (high risk)** | 469 (32.3) | 425 (34.2) | 831 (25.6) | 742 (7.8) | 2,467 (15.9) |
| Chemotherapy-treated cancer | 189 (40.3) | 186 (43.8) | 408 (49.1) | 388 (52.3) | 1,171 (47.5) |
| Hematopoietic stem cell transplant | 148 (31.6) | 127 (29.9) | 191 (23.0) | 10 (1.3) | 476 (19.3) |
| Solid organ transplant | 120 (25.6) | 109 (25.6) | 168 (20.2) | 74 (10.0) | 471 (19.1) |
| Hereditary Immune deficits | 75 (16.0) | 63 (14.8) | 122 (14.7) | 108 (14.6) | 368 (14.9) |
| Chronic autoimmune or inflammatory diseases | 45 (9.6) | 41 (9.6) | 92 (11.1) | 158 (21.3) | 336 (13.6) |
| HIV | 62 (13.2) | 26 (6.1) | 30 (3.6) | 16 (2.2) | 134 (5.4) |
| Asplenia or hyposplenia | ≤10 | <10 | <10 | <10 | 20 (0.8) |
| **Non immunocompromised patients with predisposing underlying disease (medium risk)** | 782 (53.8) | 673 (54.2) | 2,064 (63.6) | 7,559 (79.0) | 11,078 (71.4) |
| Chronic respiratory disease | 609 (77.9) | 523 (77.7) | 1,459 (70.7) | 3,587 (47.5) | 6,178 (55.8) |
| Heart failure | 149 (19.1) | 180 (26.7) | 785 (38.0) | 4,687 (62.0) | 5,801 (52.4) |
| Diabetes | 185 (23.7) | 237 (35.2) | 843 (40.8) | 2,482 (32.8) | 3,747 (33.8) |
| Chronic coronary disease | 82 (10.5) | 105 (15.6) | 458 (22.2) | 1,803 (23.9) | 2,448 (22.1) |
| Renal disease | 40 (5.1) | 57 (8.5) | 276 (13.4) | 1,953 (25.8) | 2,326 (21.0) |
| Valvular disease | 53 (6.8) | 47 (7.0) | 256 (12.4) | 1,596 (21.1) | 1,952 (17.6) |
| Stroke | 32 (4.1) | 28 (4.2) | 106 (5.1) | 622 (8.2) | 788 (7.1) |
| Chronic liver disease | 83 (10.6) | 46 (6.8) | 133 (6.4) | 195 (2.6) | 457 (4.1) |
| Obesity | 21 (2.7) | 17 (2.5) | 30 (1.5) | 38 (0.5) | 106 (1.0) |
| Cyanotic heart disease | 12 (1.5) | <10 | 18 (0.9) | 18 (0.2) | 54 (0.5) |
| **Other comorbidities** | 50 (3.4) | 45 (3.6) | 105 (3.2) | 551 (5.8) | 751 (4.8) |
| Alzheimer's disease | <10 | <10 | <10 | 237 (43.0) | 248 (33.0) |
| Dementia (other than Alzheimer) | <10 | <10 | 24 (22.9) | 197 (35.8) | 224 (29.8) |
| Parkinson's disease | <10 | <10 | 28 (26.7) | 104 (18.9) | 135 (18.0) |
| Tobacco use | 29 (58.0) | 23 (51.1) | 29 (27.6) | 30 (5.4) | 111 (14.8) |
| Alcohol use | 18 (36.0) | 13 (28.9) | 26 (24.8) | 40 (7.3) | 97 (12.9) |
| Sleep apnoea | 10 (20.0) | 13 (28.9) | 23 (21.9) | 39 (7.1) | 85 (11.3) |
| **Mean number of comorbidities, among those cited above (SD)** | 2.6 (1.8) | 2.8 (1.8) | 2.7 (1.8) | 2.5 (1.6) | 2.6 (1.7) |

### Description of the index hospitalisations

Across all years, the main diagnosis for hospitalisation was *pneumonia due to RSV* (ICD10 J12.1), which was recorded either as the principal reason for hospitalisation (32.4% of cases) or as an associated diagnosis (26.5% of cases) (Table S4, Supplementary materials). The second most common diagnosis was *acute bronchitis due to RSV* (ICD10 J20.5) as principal reason for hospitalisation (15.8%) or associated diagnosis (16.2%). Over three quarters of index hospitalisations were reported following an emergency department visit (76.5%). One quarter (25.4%) required ICU admission and 4.5% included admission to SSR follow-up and rehabilitation care with RSV infection (Table S2, Supplementary materials).

Median hospital stay was 11 days (Q1; Q3: 7.0; 17.0). Patients over 75 years had the longest hospital stays (median 11.0 days, Q1; Q3: 7.0-18.0), but were least likely to be admitted to ICU during hospitalisation (18.5%) (Table S2). Median length of stay was 10.0 days for patients aged 65 to 74 years, 10.0 days for patients 60-64 years and 9.0 days for patients aged 50 to 59 years (Table S2). Median length of hospitalisation was 10.0 days for high-risk patients, compared to 11.0 for medium risk, 10.0 for other risks and 8.0 for no comorbidities. Hospitalisations for high and medium risk patients were more likely to require ICU (27.9% and 27.4% respectively), compared to 9.3% for other risk patients and 12.3% for patients with no comorbidities.

### Mortality and rehospitalisations

Hospitalisation with RSV infection ended with death for 8.5% of patients overall (Table 3). An additional 3.8% of patients overall died within 30 days of discharge (at home or during rehospitalisation). The highest mortality rates occurred for patients 75 years and over (10.1% during hospitalisation and an additional 4.8% within 30 days) and for patients aged 65-74 years (7.1% during hospitalisation and an additional 2.5% within 30 days) (Table 3). High-, medium- and other risk patients were more likely to die during their hospitalisation than patients with no comorbidities (8.5%, 9.0%, 7.9% *vs.* 4.5%, respectively) (Table S5). Compared to patients with no comorbidities, mortality rates at 30 days were also higher for high-, medium- and other risk patients (4.3%, 3.9% and 5.9% *vs.* 1.3% respectively) (Table S5 supplementary material).

**Table 3.** In-hospital and short-term mortality, rehospitalisation and nursing home admission for patients hospitalised for respiratory syncytial virus in France between 2015 and 2022 by age group.

|  | 50 to 59 years<br>N=1,453 (%) | 60 to 64 years<br>N=1,242 (%) | 65 to 74 years<br>N=3,244 (%) | 75 years or<br>over<br>N=9,570 (%) | All patients<br>(50y+)<br>N=15,509 (%) |
| --- | --- | --- | --- | --- | --- |
| **In-hospital death** | 66 (4.5) | 68 (5.5) | 230 (7.1) | 962 (10.1) | 1,326 (8.5) |
| **Patients still alive at hospital discharge, N** | 1,387 | 1,174 | 3,014 | 8,608 | 14,183 |
| Mortality rate |  |  |  |  |  |
| 30 days | 26 (1.9) | 27 (2.3) | 75 (2.5) | 414 (4.8) | 542 (3.8) |
| 90 days | 46 (3.3) | 57 (4.9) | 174 (5.8) | 859 (10.0) | 1,136 (8.0) |
| Overnight rehospitalisation for all causes |  |  |  |  |  |
| 30 days | 258 (18.6) | 267 (22.7) | 583 (19.3) | 1,413 (16.4) | 2,521 (17.8) |
| 90 days | 452 (32.6) | 424 (36.1) | 1,063 (35.3) | 2,598 (30.2) | 4,537 (32.0) |
| Overnight rehospitalisation for respiratory causes<sup>a</sup> |  |  |  |  |  |
| 30 days | 73 (5.3) | 74 (6.3) | 150 (5.0) | 363 (4.2) | 660 (4.7) |
| 90 days | 142 (10.2) | 124 (10.6) | 317 (10.5) | 675 (7.8) | 1,258 (8.9) |
| Overnight rehospitalisation for cardiorespiratory causes<sup>b</sup> |  |  |  |  |  |
| 30 days | 90 (6.5) | 105 (8.9) | 235 (7.8) | 676 (7.9) | 1,106 (7.8) |
| 90 days | 191 (13.8) | 176 (15.0) | 493 (16.4) | 1,314 (15.3) | 2,174 (15.3) |
| Admission to a nursing home |  |  |  |  |  |
| 30 days | 0 (0.0) | <10 | <10 | 101 (1.2) | 104 (0.7) |
| 90 days | <10 | <10 | 10 (0.3) | 269 (3.1) | 281 (2.0) |

<sup>a</sup>ICD10 codes starting with J
<sup>b</sup>ICD10 codes starting with I or J. In these analyses, only hospitalisation that included an overnight stay were counted.

Following discharge from the index RSV hospitalization, 2,521 patients (17.8%) were rehospitalised within 30 days for all causes, among which, 1,106 patients (43.9%) were rehospitalised for cardiorespiratory causes (Table 3). Required rehospitalisations varied across age groups, from 16.4% for 75 years and over to 22.7% for 60 to 64 years. High-risk patients were much more likely to require rehospitalisation than any other risk group of patients (27.9% *vs*. 16.7%, 12.0% and 11.1% for medium, other and no risk groups, respectively) (Table S5).

### Healthcare use (outside the index hospitalisation)

The proportions of patients seeking GP appointments increased from 52.1% in the pre-RSV infection period, to 68.4% in the pre-hospitalisation period, and 60.1% post-hospitalisation (Table 4). The largest change in healthcare use was an increase in X-rays of the respiratory system, from 8.6% of patients receiving imaging before infection, 64.2% receiving imaging during the period of infection and 15.7% of patients in the 30-day post-hospitalisation period (Table 4). Pre-hospitalisation, the most common treatments were respiratory system treatments (33.4%), analgesics/antipyretics (painkillers) (32.4%), and antibiotics (30.0%), and patterns were similar across age groups. Post-hospitalisation, the use of all three of these treatments was greater than pre-infection (Table 4).

**Table 4.** - Healthcare use before RSV infection and before and after index hospitalisation for patients hospitalised with RSV infection in France between 2015 and 2022 (and alive at discharge) by age group

|  | 50 to 59<br>N=1,387 (%) | 60 to 64<br>N=1,174 (%) | 65 to 74<br>N=3,014 (%) | 75 years or over<br>N=8,608 (%) | All patients<br>N=14,183 |
| --- | --- | --- | --- | --- | --- |
| **General practitioner visit** |  |  |  |  |  |
| Pre-infection<sup>a</sup> | 681 (46.9) | 559 (45.0) | 1678 (51.7) | 5161 (53.9) | 8079 (52.1) |
| Pre-hospitalisation | 945 (65.0) | 780 (62.8) | 2215 (68.3) | 6665 (69.6) | 10605 (68.4) |
| Post-hospitalisation | 820 (59.1) | 676 (57.6) | 1768 (58.7) | 5253 (61.0) | 8517 (60.1) |
| **Specialist visit** |  |  |  |  |  |
| Pre-infection | 224 (15.4) | 257 (20.7) | 583 (18.0) | 1320 (13.8) | 2384 (15.4) |
| Pre-hospitalisation | 286 (19.7) | 252 (20.3) | 620 (19.1) | 1485 (15.5) | 2643 (17.0) |
| Post-hospitalisation | 242 (17.4) | 213 (18.1) | 550 (18.2) | 1153 (13.4) | 2158 (15.2) |
| **X-ray of respiratory system<sup>b</sup>** |  |  |  |  |  |
| Pre-infection | 152 (11.0) | 123 (10.5) | 291 (9.7) | 656 (7.6) | 1,222 (8.6) |
| Pre-hospitalisation | 837 (60.3) | 752 (64.1) | 1,927 (63.9) | 5,586 (64.9) | 9,102 (64.2) |
| Post-hospitalisation | 259 (18.7) | 246 (21.0) | 560 (18.6) | 1,165 (13.5) | 2,230 (15.7) |
| **All laboratory tests** |  |  |  |  |  |
| Pre-infection | 434 (31.3) | 430 (36.6) | 1,234 (40.9) | 3,344 (38.8) | 5,442 (38.4) |
| Pre-hospitalisation | 526 (37.9) | 494 (42.1) | 1,335 (44.3) | 3,501 (40.7) | 5,856 (41.3) |
| Post-hospitalisation | 589 (42.5) | 536 (45.7) | 1,467 (48.7) | 4,039 (46.9) | 6,631 (46.8) |
| **Respiratory treatment<sup>c</sup>** |  |  |  |  |  |
| Pre-infection | 409 (29.5) | 393 (33.5) | 1,029 (34.1) | 2,209 (25.7) | 4,040 (28.5) |
| Pre-hospitalisation | 526 (37.9) | 455 (38.8) | 1,142 (37.9) | 2,620 (30.4) | 4,743 (33.4) |
| Post-hospitalisation | 556 (38.3) | 454 (36.6) | 1,178 (36.3) | 2,463 (25.7) | 4,651 (30.0) |
| **Antiviral medication<sup>d</sup>** |  |  |  |  |  |
| Pre-infection | 151 (10.9) | 145 (12.4) | 235 (7.8) | 173 (2.0) | 704 (5.0) |
| Pre-hospitalisation | 152 (11.0) | 134 (11.4) | 208 (6.9) | 162 (1.9) | 656 (4.6) |
| Post-hospitalisation | 190 (13.1) | 160 (12.9) | 241 (7.4) | 217 (2.3) | 808 (5.2) |
| **Pain-killers<sup>e</sup>** |  |  |  |  |  |
| Pre-infection | 336 (24.2) | 319 (27.2) | 834 (27.7) | 2,824 (32.8) | 4,313 (30.4) |
| Pre-hospitalisation | 448 (32.3) | 359 (30.6) | 966 (32.1) | 2,828 (32.9) | 4,601 (32.4) |
| Post-hospitalisation | 413 (28.4) | 354 (28.5) | 923 (28.5) | 3,042 (31.8) | 4,732 (30.5) |
| **Antibiotics<sup>f</sup>** |  |  |  |  |  |
| Pre-infection | 333 (24.0) | 293 (25.0) | 634 (21.0) | 1,186 (13.8) | 2,446 (17.2) |
| Pre-hospitalisation | 510 (36.8) | 415 (35.3) | 968 (32.1) | 2,364 (27.5) | 4,257 (30.0) |
| Post-hospitalisation | 483 (33.2) | 420 (33.8) | 928 (28.6) | 1,892 (19.8) | 3,723 (24.0) |

<sup>a</sup>Pre-infection = From 60 days before to 30 days before RSV hospitalisation [-60 days; - 30 days]. Pre-hospitalisation = 30 days before RSV hospitalisation; Post-hospitalisation = 30 days after discharge. <sup>b</sup>As defined by French CCAM codes: GEQH001 – Bronchographie (bronchography), LIQK002 - Radiographie du thorax avec radiographie du squelette du thorax (Chest X-ray with chest skeleton X-ray), ZBQK002 - Radiographie du thorax (Chest X-ray), ZBQK003 - Examen radiologique dynamique du thorax, pour étude de la fonction respiratoire et/ou cardiaque (Dynamic chest X-ray examination, for study of respiratory and/or cardiac function). <sup>c</sup>Anatomical therapeutic chemical (ATC) classification beginning with 'R'. <sup>d</sup>ATC codes beginning with J05. <sup>e</sup>AINS M01A, Paracetamol N02BE01, or Aspirin N02BA01. <sup>f</sup>ATC Codes J01.

### Hospitalisation and healthcare costs

The average direct cost of index RSV hospitalisation was €6,876 (SD €9,346), with the highest costs observed for 60 to 64-year-old patients (mean €9,275, SD=€14,514) and the lowest costs for patients 75 years and over (mean €5,854, SD=€6,178) (Table 5). Index hospitalisation costs were lowest overall for patients with no identified comorbidity risks (mean €5,087, SD=€7,512) and highest for high-risk immunocompromised patients (mean €9,618, SD=€15,070) (Table S6 supplementary materials).

**Table 5.** Index hospitalisation cost and 30-day global healthcare costs before respiratory syncytial virus (RSV) infection and before and after RSV hospitalisation for patients in France hospitalised with RSV between 2015 and 2022 by age group

|  | 50 to 59 years<br>N=1,453 | 60 to 64 years<br>N=1,242 | 65 to 74 years<br>N=3,244 | 75 years or over<br>N=9,570 | All patients (50y+)<br>N=15,509 |
| --- | --- | --- | --- | --- | --- |
| **Cost of index hospitalisation in euros<sup>a</sup>, N** | 1,422 | 1,210 | 3,172 | 9,365 | 15,169 |
| Mean (SD) | 8,729 (13,274) | 9,275 (14,514) | 8,149 (11,721) | 5,854 (6,178) | 6,876 (9,346) |
| Median (Q1; Q3) | 4,435 (2,752; 8,615) | 4,594 (2,899; 9,880) | 4,395 (2,945; 8,787) | 4,180 (3,133; 6,251) | 4,252 (3,077; 7,007) |
| [Min; Max] | [268;135,905] | [275;157,393] | [259;252,786] | [279;188,498] | [259;252,786] |
| **Healthcare costs by period, in euros** |  |  |  |  |  |
| **Pre-infection: 30 days before RSV infection<sup>b</sup>** |  |  |  |  |  |
| Mean (SD) | 2,853 (5,678) | 3,157 (7,331) | 2,650 (8,217) | 1,567 (3,514) | 2,041 (5,426) |
| Median (Q1; Q3) | 650 (87; 2,761) | 745 (138; 3,181) | 524 (149; 2,356) | 387 (135; 1,115) | 436 (132; 1,490) |
| [Min;Max] | [0;50,214] | [0;95,620] | [0;359,717] | [0;57,398] | [0;359,717] |
| **Pre-hospitalisation: 30 days prior to RSV hospitalisation** |  |  |  |  |  |
| Mean (SD) | 2,997 (5,064) | 3,073 (5,258) | 2,485 (4,914) | 1,694 (3,093) | 2,092 (3,981) |
| Median (Q1; Q3) | 958 (181; 3,514) | 1,035 (214; 3,519) | 708 (220; 2,765) | 558 (197; 1,564) | 634 (201; 2,078) |
| [Min;Max] | [0;54,046] | [0;53,395] | [0;129,619] | [0;39,392] | [0;129,619] |
| **Post-hospitalisation: 30 days following RSV hospitalisation<sup>a</sup>, N** | 1,387 | 1,174 | 3,014 | 8,608 | 14,183 |
| Mean (SD) | 5,211 (10,453) | 5,554 (12,067) | 4,574 (8,907) | 3,945 (8,726) | 4,336 (9,278) |
| Median (Q1; Q3) | 1,670 (498; 5,333) | 1,857 (514; 5,912) | 1,288 (436; 5,260) | 932 (358; 4,163) | 1,117 (389; 4,700) |
| [Min;Max] | [0;138,828] | [0;192,345] | [0;151,688] | [0;192,774] | [0;192,774] |
| **Additional healthcare costs post RSV hospitalisation<sup>a,c</sup>, N** | 1,387 | 1,174 | 3,014 | 8,608 | 14,183 |
| Mean (SD) | 2,477 (10,010) | 2,502 (11,661) | 2,009 (11,042) | 2,440 (8,715) | 2,357 (9,648) |
| Median (Q1; Q3) | 459 (8; 2,321) | 401 (-11; 2,354) | 344 (-34; 2,292) | 298 (2; 2,254) | 323 (0; 2,292) |
| [Min;Max] | [-84,267;192,345] | [-357,893;151,325] | [-43,349;164,849] | [-357,893;192,345] | [-357,893;164,849] |

<sup>a</sup>Data for all patients alive at hospital discharge; <sup>b</sup>Pre-infection period: Before RSV infection – From two months before to one month before hospitalisation [-60 days; - 30 days ]. <sup>c</sup> Additional healthcare costs are the difference between the post-RSV hospitalisation 30-day costs and the 30-day pre-hospitalisation costs and thus could be less than zero. These costs are considered attributable largely to RSV. N are given in column headings, unless otherwise specified. SD – standard deviation, Q1; Q3 - First and third quartiles

Average healthcare costs increased substantially across the three defined time periods. During the pre-infection reference period, mean costs were (€2,041, SD =€5,426); during the pre-hospitalisation period, mean costs were €2,092, SD=€3,981); and during the post-hospitalisation 30-day period, they were (€4,336, SD=€9,278) (Table 5). This represented an average crude additional per patient cost of €2,357 (SD=€9,648) in our population. There did not appear to be any age-based differences. There was a large range in additional post-hospitalisation costs, with maximum additional costs over 30 days exceeding €190,000 for one patient.

The least square means obtained from the adjusted linear mixed model (Table S7, Figure 2) show additional cost variability across patient groups (age, sex, year and comorbidity) compared to a reference male patient of 50-59 years without comorbidities, hospitalised for RSV during the 2019-2020 season incurring costs of €2,786. All profiles of patients alive at discharge had higher healthcare expenditures in the 30 days post-hospitalisation than in the 30 days prior to infection. The multivariate model showed little variation between age groups. There were no interactions between age and risk groups. Patients with no identified comorbidities had the highest additional cost burden due to high post-hospitalisation costs (mean €3,245, SD=€11,204) in a population with initially small healthcare expenditures during the reference period (mean €655, SD=€2,062) (Table S5).

**Figure 2.**
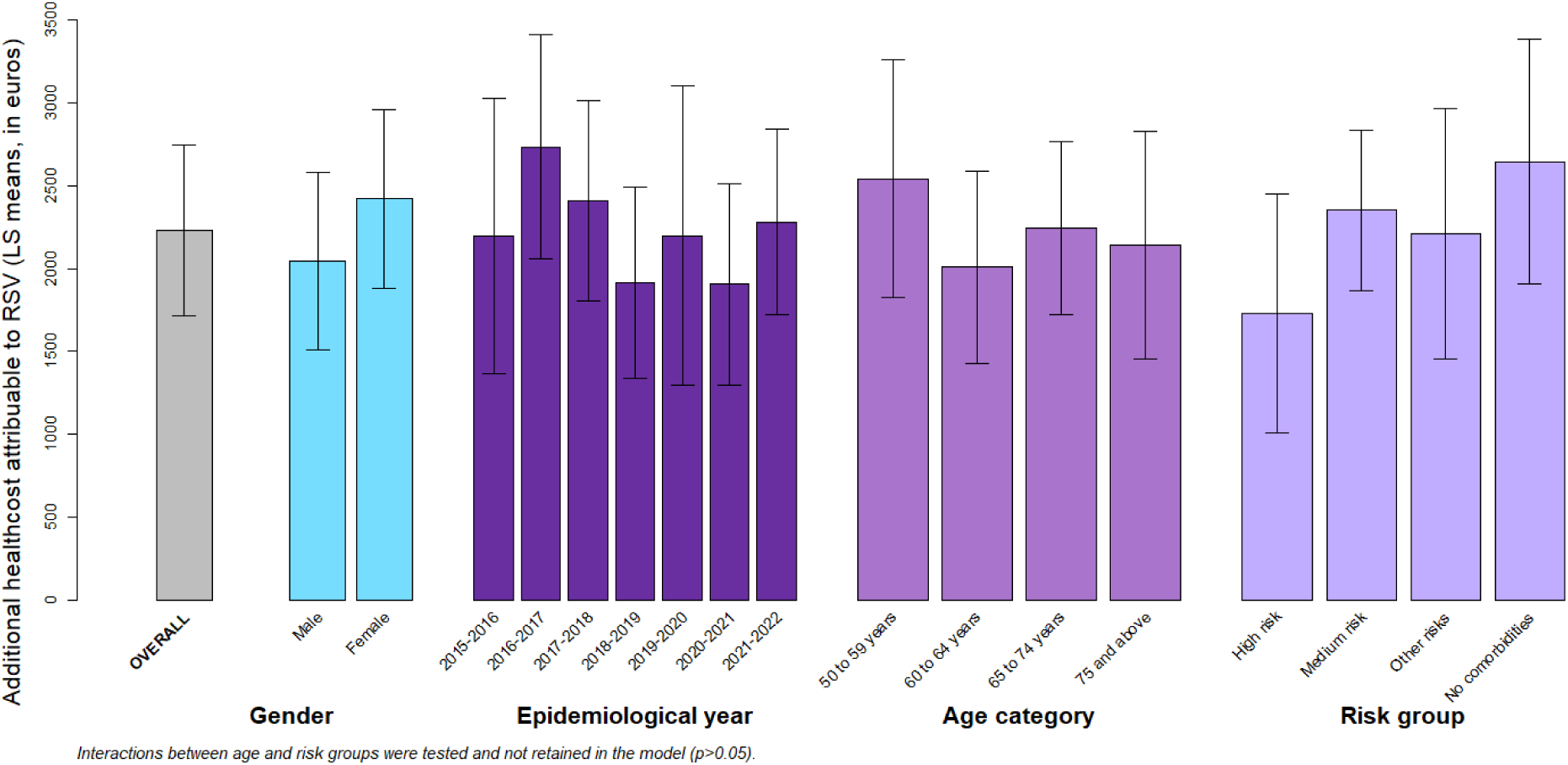
Least-square means of additional costs attributed to RSV by risk factor, estimated by a linear mixed effects model.

Re-hospitalisations within 30 days were infrequent (4.7% of all patients alive at discharge, Table 3), but costly and they accounted for a large proportion of the additional costs. Inpatient costs increased abruptly at an additional €2,059 (SD=€9,618) over 30 days, compared with an increase of €283 (SD €1,551) in outpatient costs (Table S8).

## Discussion

By combining hospital discharge database analysis with outpatient healthcare costs from the community at a national level, our study presents a comprehensive description of RSV patients, hospitalisation and costs. Our analysis of seven epidemiological years has revealed the significant hospitalisation and economic burden of RSV for adults over 50 years in France. During this period, there were over 15,000 hospitalisations, resulting in 1,326 deaths. Additionally, all surviving patients, irrespective of age or risk profile, faced increased healthcare costs. Most patients who required hospitalisation were immune-compromised or had underlying comorbidities and /or conditions (92.2%), and > 82% of hospitalisations occurred for patients aged 65 years and above (61.7% of hospitalisations occurred for patients aged 75 years and above). RSV infection thus appears to be a trigger for hospitalisation specifically for two populations: patients aged 65 years and over, irrespective of immune-status and presence of comorbidities, or patients aged 50 to 64 years who are immune-compromised (one third of them) or who have a chronic condition (half of them).

With 1,326 deaths recorded over the study period, the 8.5% rate of in-hospital mortality observed in our study was similar to the 7.3% reported previously in France (11) and the 8% reported by Falsey *et al.* in the United States (5) and within the range reported recently for patients aged 60 and over (from 6.73% to 10.14% depending on country)(26). However, these rates are higher than previous estimates ranging from 1.6% (95% CI: 0.7–3.8) (27) to 4.9% (7), 6.1% (95% CI: 3.3–11.0) (15) and 7.1% (95% CI: 5.4–9.3) (28) and may reflect the greater severity of RSV infection in the studied population, or differences in healthcare systems and access to care. Our observed rate is closer to the meta-estimate for patients with comorbidities at 11.7% in-hospital fatality rate, as reported by a recent systematic review of industrialised countries (29). Nonetheless, the present results confirm that in-hospital mortality rates increase with comorbidity risk and with age (30). For comparison, country-specific influenza mortality rates in 2010-2018 ranged from 4.1 to 9.3% in the adult population 50 years and over (31).

The severity of infection is shown by the high ICU admission rate at 25.4% of patients across all age and risk groups. In the United States, for RSV hospitalisations for patients of the same age (50+) across five seasons, Woodruff *et al.* recently reported that 18.6% of patients required ICU admission (7) and other international studies have estimated ICU admissions between 15 to 16.5% (32,33). A previous analysis of French hospital claims data reported that 10.4% to 19.3% of patients needed admission to the ICU, depending on the definition used (11). Similar to Nuttens *et al.* (12), the younger patients in our study were likely to require ICU (37.9% for 50-59 years), primarily because of the severity of comorbidities observed in this population.

At €6,876, the average RSV hospitalisation cost varied across age-groups, with lower costs for patients 75 and older (€5,854) and higher costs for patients 60-64 years (€9,275). Immune-compromised patients had the highest costs overall (€9,618) and patients with no risks had the lowest costs (€5,087). RSV hospitalisation costs reported in the literature for the United States have ranged substantially, from as low as $8,049 (34) to as high as $58,117 (35). However, the hospitalisation costs we observed are within the range of the costs previously reported in France : €6,252 for the 2018/19 season for all patients and €8,558 for patients with comorbidities (11), and €4,656 to €6,167 depending on age category and season (12). Hospitalisation costs observed increased with comorbidity risk or immunocompromised status, as reported in the literature (34,36,37). Previous United States and French studies have demonstrated the costs of RSV hospitalisations to be similar to the costs of influenza hospitalisation (12,38).

There are several methods for estimating the economic burden of RSV hospitalisations (see Grace *et al.* (39) for a review). Using a pre- and post-hospitalisation approach similar to Wyffels *et al*. (40) and Mesa-Frias *et al.* (41), we present data on additional healthcare costs compared with a reference period just prior to RSV infection. For a patient aged 50-59 years without comorbidities, the additional cost of RSV hospitalisation (excluding costs of the index hospitalisation) compared with a reference period was estimated at €2,787 (Table S7). The additional cost of RSV infection was high for all patients; however, the costs were relatively higher for patients without comorbidities because these patients consumed less during the reference period. This aligns with earlier findings reporting a 35% increase in outpatient costs for low-risk RSV patients compared to an 11% increase for high-risk RSV patients (40). Immunocompromised patients or patients with high-risk comorbidities consumed more healthcare at baseline and during all subsequent periods (Table S5).

When we look at the breakdown of additional costs between inpatient and outpatient costs, the average additional amount of €2,357 per patient (Table S6) is largely explained by the additional inpatient rehospitalisation costs observed (€2,059, Table S8). As only 17.8% of patients required rehospitalisation, the impact of this small proportion of patients with extreme costs should be taken into account for generalisability of results.

Given the availability of prophylaxis for RSV, these results should be considered in the context of current vaccination recommendations. The current recommendations in France, published in July 2024, are to vaccinate adults over 75 years of age and those 65 years and over with chronic respiratory or cardiac pathologies (42). With 21% of the hospitalised population aged between 65-74 years, and over 80% of patients under 65 years either having underlying predispositions (53.8% and 54.2%, respectively) or being immune-compromised (32.3% and 34.2% respectively), together with median hospitalization costs exceeding €9,000 in patients aged 65-74 or immune-compromised patients, these findings highlight the need to consider broadening vaccination recommendations to mitigate this considerable healthcare and economic burden. Vaccination and prevention of RSV infections in people between 50 and 65 years of age with comorbidities can also reduce indirect costs (e.g., work days lost) (10) because a large proportion of this population is still active in the workforce, though this was out of scope of the present study.

While using the national SNDS database is a strength of this study, it also has its limits, for example laboratory test results are not included and the regulatory processes to obtain access to data result in delays before results can be analysed and published. In addition, limitations of the hospitalisation context should be taken into account for interpretation, as only the most severe cases of RSV require hospitalisation. Due to the absence of RSV testing, low diagnostic sensitivity and misuse of RSV-specific ICD-10 codes (15,43–47), our study does not estimate incidence. A recent systematic literature review (SLR) indicated that conventional laboratory methods detect only about 62% of adult RSV infections, meaning that over one third of cases may be missed unless more sensitive molecular diagnostics are used (48). Hospitalisations for RSV for adults in France have been modelled in a recent study by Nuttens *et al.* (12), finding 15 times more hospitalisations using a modelling approach compared to identifying hospitalisations by ICD-10 code. Changes in the testing protocol should also be considered, which may explain variations in the number of cases and additional costs over time (11,49). This was also observed in an earlier French study (11) and recent research has indicated an increase in PCR testing for RSV over the period 2017-2023 (22). Comparison between laboratory tests and hospitalisation codes shows that RSV is still under-coded even when diagnostic tests are conducted (22). Finally, for outcomes within 90 days following discharge from RSV hospitalisation, the link with index hospitalisation is open to discussion, especially as influenza epidemics tend to follow RSV epidemics every year.

## Conclusions

Our analysis of seven years of healthcare and hospital claims data showed that RSV leads to a significant hospitalization burden, including prolonged stays, high ICU admission rates, and increased post-hospitalization costs, particularly for patients aged 50+ with comorbidities and patients aged 65 years and above. Broader vaccination prevention strategies, including patients with underlying conditions and/or immunosuppression, as well as lower age thresholds, could help to reduce the burden of RSV in adults. This study not only describes the current burden, it also provides essential data for future modelling of the impact of vaccination.

## Supporting information

Vacheret et al.Sup_Mat_RESVYRII

## Data Availability

The data used to support the conclusions of this article are available from the French National Health Data System (SNDS)

## Declarations

### Ethics approval and consent to participate

This study was carried out in accordance with French medical data privacy laws and obtained the required ethics approval for use of the SNDS by the *Comité d’Expertise pour les Recherches, les Etudes et les Evaluations dans le domaine de la Santé* (CESREES) and the *Commission Nationale de l’Informatique et des Libertés* (CNIL).

### Consent for publication

Not applicable

### Author contributions

MV, PL contributed to the study conception and design, as well as the interpretation of data and critical review of the manuscript. JEK, SF, EB, LW, JSC, HL, PV, LB, CBF, LI contributed to the methodology and data analysis. MV, JEK, SF, EB, PL contributed to the study design, and interpretation of data. All authors reviewed and approved the final manuscript ahead of submission.

## Acknowledgements

The authors are grateful to the Cellule de la CNAM en Charge de l’accompagnement des Demandes D’extraction (DEMEX) teams, especially Julien Brand and Marjorie Boussac, at the Caisse Nationale de l’Assurance Maladie (CNAM) for their assistance with data extraction.

## List of abbreviations

COPD: Chronic Obstructive Pulmonary Disease
DCIR: National claims database for healthcare use in France (Datamart de Consommation InterRegimes)
GP: General Practitioner
HIV: Human Immunodeficiency Virus
ICD10: International Classification of Diseases, 10th Revision
ICU: Intensive Care Unit
MSO: Medical Surgery and Obstetrics
PMSI: Programme de Médicalisation des Systèmes d’Information (French hospital discharge data)
RSV: Respiratory Syncytial Virus
SD: Standard Deviation
SLR: Systematic Literature Review
SMR: Soins Médicaux et de Réadaptation (Medical Rehabilitation Care)
SNDS: Système National des Données de Santé (French National Health Database)
SSR: Soins de Suite et de Réadaptation (Follow-up and Rehabilitation Care)

