## Supplementary material for "Hospitalisation and healthcare burden of respiratory syncytial virus in adults over 50 years in France": Vacheret et al.Sup_Mat_RESVYRII

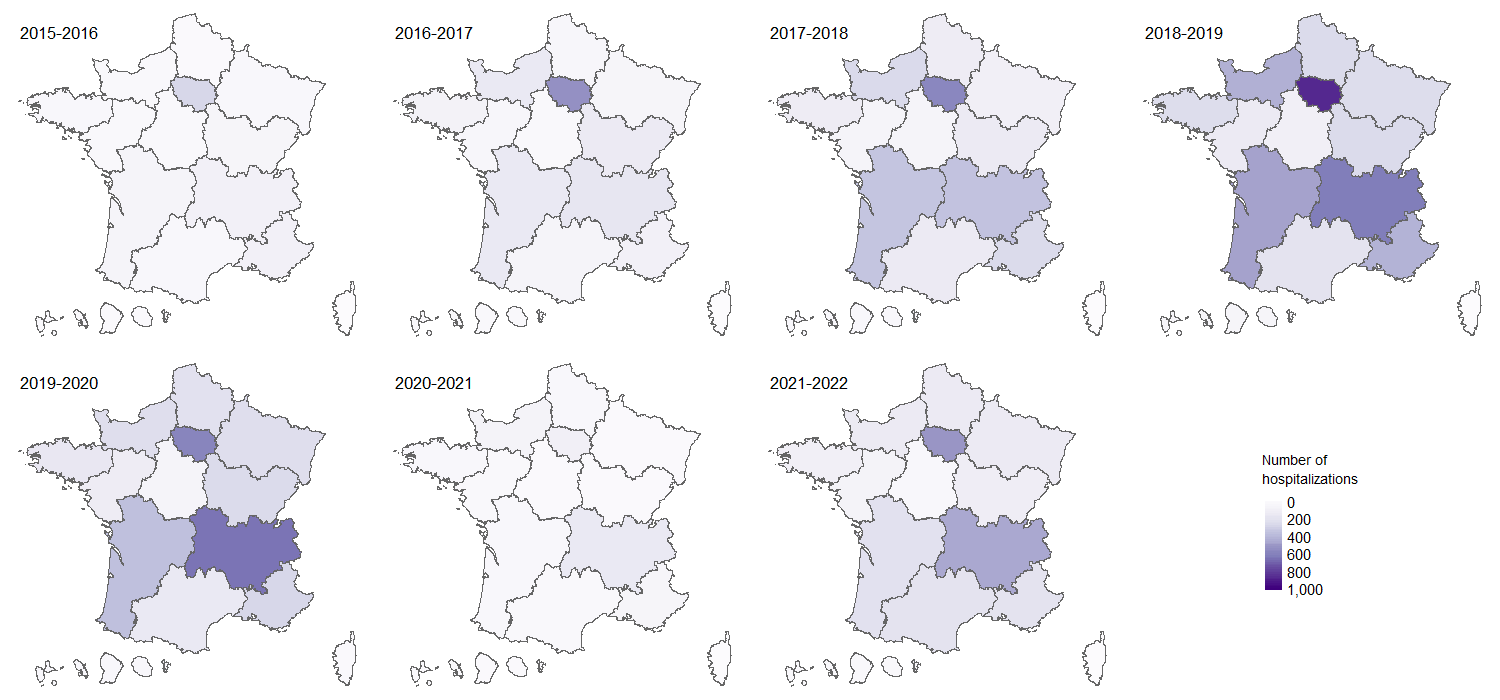

Figure S1**:** Regional map of the number respiratory syncytial virus (RSV) hospitalizations (identified by hospital regional code) by epidemiological year and French region.

Table S1: Algorithms used for comorbidity identification in the French national healthcare claims database, SNDS, derived from the Méthodologie médicale de la cartographie des pathologies et des dépenses, version G11 (CNAM)

| **Comorbidity** | **Algorithm** | **LTD** | | **Hospitalization and procedures** | | | | **Drugs, devices and lab tests** | |
| --- | --- | --- | --- | --- | --- | --- | --- | --- | --- |
|  |  | **ICD-10** | **Years** | **Diagnoses (ICD-10) or GHM** | **Diagnoses** | **Procedures (CCAM)** | **Years** | **Codes (ATC, LPP, TNB)** | **Years** |
| **Asplenia or hyposplenia** | LTD and/or Hospitalizations and/or procedures | D730 | n-1 | C261, D561, D570, D572, D730-D735, D738, D739, P151, Q890, S3600, S3601, Z8502 | PD / RD / SAD ‡ | FFFA001, FFFA002, FFFC001, FFFC420, FFQX005, HEPA004, HEPA007, HNFA004, HNFA006, HNFA010, HNFA013, HNFC002, HNQX007 | n-4 to n-1 |  |  |
| **Hereditary Immune deficits** | LTD and/or Hospitalizations | D81-D89, G113 | n-1 | D81-89, G11.3 | PD / RD / SAD |  | n-1 |  |  |
| **HIV** | LTD and/or Hospitalizations and/or Drugs and/or Biologic tests | B20*, B21*, B22*, B23*, B24*, Z21 | n-1 | B20*, B21*, B22*, B23*, B24*, Z21, F024, Z206 | MCO: PD / RD |  | n-4 to n-1 | Drugs: ≥3 reverse, protease transcriptase inhibitors and HIV antivirals in a year  Lab tests (TNB): 0805, 0806, 1691, 4117, 4122 | n-1 |
|  |  |  |  | R75, O987 | PSY: PD / AD |  |  |  |  |
|  |  |  |  | B20*, B21*, B22*, B23*, B24*, Z21 | PD / RD (RUM) or SAD |  | n-1 |  |  |
| **Chemotherapy-treated cancer** | [3 drug dispensals AND [LTD or HOSPIT PD/RD] ]  and/or [chemotherapy as PD + cancer as DR/SAD] | C* | n-1 | Z511, Z08.2, C* | PD or DR |  | n-1 | Drugs: ≥3 oral anticancer drugs | n-1 |
| **Solid organ transplant** | LTD and/or Hospitalizations and/or Drugs | LTD28 (Following organ transplant) | n-1 | Z940-944, Z9481, Z9482 SEL_CODE_RESV_SORGT | PD / RD / SAD | JAEA003, JAEA002, DZEA002, DZEA003, DZEA001, DZEA004, GFEA005, GFEA002, GFEA003, GFEA007, GFEA004, GFEA006, HGEA002, HGEA004, HLEA001, HNEA900, HNEA002, HNEH900 | n-4 to n-1 | Drugs: ≥3 anti-rejection drugs in a year | n-1 |
| **Hematopoietic stem cell transplant** | Hospitalisation and/or GHM and/or procedure |  |  | Z948* | PD / RD / SAD | FELF010, FELF009 | n-4 to n-1 |  |  |
|  |  |  |  | DRG: 27Z02*, 27Z04J |  |  |  |  |  |
| **Chronic autoimmune or inflammatory diseases treated by immunosuppressive or biologic drugs** | [LTD and/or Hospitalizations] AND Drugs | K50*, K51*, L93*, L94*, M05*, M06*, M45*, M46*, M074, M075, M30*, M08, M09, M31*, M32*, M33*, M34*, M35*, M36 | n-1 | K50*, K51*, L93*, L94*, M05*, M06*, M45*, M46*, M074, M075, M30*, M08, M09, M31*, M32*, M33*, M34*, M35*, M36 | PD / RD |  | n-4 to n-1 | Drugs: ≥3 immunosuppressive or biologic drugs in a year | n-2 to n-1 |
|  |  |  |  |  | SAD |  | n-1 |  |  |
| **Nephrotic Syndrome** | LTD and/or Hospitalizations | N04* | n-1 | N04* | PD / RD / SAD |  | n-4 to n-1 |  |  |
| **Chronic respiratory disease** | LTD and/or Hospitalizations | J40*, J41*, | n-1 | J40*, J41*, J42*, J43*, J44*, J45*, J46*, J47*, J96*, J98* with exclusion of J960* and J969* | PD / RD |  | n-4 to n-1 | Drugs: ≥3 treatments dispensing in the year: R03 | n-1 |
|  |  | J42*, J43*, J44*, J45* J46*, J47*, J96*, J98* |  |  | SAD |  | n-1 |  |  |
| **Cyanotic heart disease** | Hospitalizations |  |  | Q20*-Q25* | PD / RD / SAD |  | n-4 to n-1 |  |  |
| **Heart failure** | LTD and/or Hospitalizations | I50*, I11*, I13* | n-1 | HF: I50* | PD (RUM) or RD |  | n-4 to n-1 |  |  |
|  |  |  |  | Complications: I110, I130, I132, I139, K761*, J81 | PD (RUM) with DR / SAD of HF |  | n-4 to n-1 |  |  |
|  |  |  |  | HF: I50* | SAD, or RD (RUM) |  | n-4 to n-1 |  |  |
| **Chronic coronary disease** | LTD and/or Hospitalizations | I20*, I21*, I22*, I23*, I24*, I25* SEL_CODE_RESV_CCD | n-1 | I20, I21, I22, I23, I24, I25 | PD (RUM) or RD |  | n-4 to n-1 |  |  |
|  |  |  |  | Complications: I25 | PD / RD / SAD |  | n-1 |  |  |
| **Valvular disease** | LTD and/or Hospitalizations | I05*, I06*, I07*, I08*, I34*, I35*, I36*, I37*, I38*, I39* | n-1 | I05*, I06*, I07*, I08*, I34*, I35*, I36*, I37*, I38*, I39* | PD / RD / SAD |  | n-4 to n-1 |  |  |
| **Stroke** | LTD and/or Hospitalizations | I60*, I61*, I62*, I63*, I64*, I67*, I68*, I69*, G81* | n-1 | I60*, I61*, I62*, I63*, I64*, I67*, I68*, I69* | PD (RUM) or RD |  | n-4 to n-1 |  |  |
|  |  |  |  | Complications: I60*, I61*, I62*, I63*, I64*, I67*, I68*, I69*  And with exclusion of I65* et I66* | PD / RD (RUM) / SAD |  | n |  |  |
| **Renal disease** | Hospitalizations | N18, Z940 |  | N18, Z940 | PD / RD / SAD |  | n-1 |  |  |
| **Chronic liver disease** | LTD and/or Hospitalizations and/or Drugs | B18*, I85*, K70*, K71*, K72*, K73*, K74* | n-1 | B18*, I85*, K70*, K71*, K72*, K73*, K74* SEL_CODE_CARTO_CLD | PD / RD |  | n-4 to n-1 | Drugs: ≥ 3 hepatitis C and/or B treatments | n-1 |
|  |  |  |  |  | SAD |  | n-1 |  |  |
| **Diabetes** | LTD and/or Hospitalizations and/or Drugs | E10*, E11*, E12*, E13*, E14* | n-1 | E10*, E11*, E12*, E13*, E14* | PD / RD |  | n-1 | Drugs: ≥3 (2 if large packaging) diabetes treatments dispensing in the yea | n-1 |
|  |  |  |  | Diabetes complications: G590, G630, G730, G990, H280, H360, I790, L97, M142*, M146*, N083* SEL_CODE_CARTO_COMP_DIAB |  |  |  |  |  |
| **Sleep apnoea** | Hospitalizations and/or device and/or procedure |  |  | G473 | PD / RD / SAD | GLMP001, LBLD017 | n-4 to n-1 | Devices: ≥2 Mandibular advancement splint (MAS) and/or continuous positive airway pressure (CPAP) | n-4 to n-1 |
| **Tobacco use** | Hospitalizations and/or services and/or drugs and/or tests |  |  | F17, | PD / RD / SAD |  | n-4 to n-1 | Drugs: ≥3 drugs used in nicotine dependence Lab tests (TNB): 9566, 9526, 9527 | n-4 to n-1 |
|  |  |  |  | Z716, |  |  |  |  |  |
|  |  |  |  | Z720 SEL_CODE_RESV_TAB |  |  |  |  |  |
| **Alcohol use** | LTD and/or hospitalizations and/or drugs | F10, K70, Z50.2, Z71.4, Z72.1 | n-4 to n-1 | F10, G31.2, G62.1, G72.1, E24.4, K86.0, T51, Z71.4, I42.6, K29.2, K70, Z50.2, Z72.1, K85.2 | PD / RD / SAD |  | n-4 to n-1 | Drugs: ≥3 drug deliveries | n-4 to n-1 |
| **Obesity** | Hospitalizations and/or procedure and/or drugs | E66 | n-1 | E66 | PD / RD | Bariatric surgery: HFCA001, HFCC003, HFFA001, HFFA011, HFFC004, HFFC018, HFGC900, HFKA001, HFKA002, HFKC001, HFMA009, HFMA010, HFMA011, HFMC006, HFMC007, HFMC008, HGCA009, HGCC027 | n-4 | Drugs: ≥3 drug deliveries of A08AB01 | n-1 |
| **Malnutrition** | Hospitalizations and/or drug |  |  | E40, E41, E42, E43, E44, E45, E46 | PD / RD |  | n-1 | Drugs: ≥3 deliveries of oral nutritional supplements | n-1 |
| **Alzheimer disease** | LTD and/or Hospitalizations and/or Drugs | F00, G30 | n-1 | F00, G30 | PD / RD |  | n-4 to n-1 | Drugs: ≥3 anti-Alzheimer’s drugs: N06DA*, N06DX01 | n-4 to n-1 |
|  |  |  |  |  | SAD |  | n-1 |  |  |
| **Parkinson disease** | LTD and/or Hospitalizations and/or Drugs | G20, F023 | n-1 | G20, F023 | PD / RD |  | n-4 to n-1 | Drugs: ≥3 anti-Parkinson drugs | n-4 to n-1 |
|  |  |  |  |  | SAD |  | n-1 |  |  |
| **Multiple sclerosis** | LTD and/or Hospitalizations and/or Drugs | G35 | n-1 | G35 | PD / RD |  | n-4 to n-1 | Drugs: ≥3 anti-multiple sclerosis drugs | n-4 to n-1 |
| **Dementia** | LTD and/or hospitalization and/or drugs (and no Alzheimer) | F00 F02 F01 F03 G30 F051 except F02.3, F0.24 | n-1 | F00 F02 F01 F03 G30 F051 except F02.3, F0.24 | MCO: DP or DR  RIP: DP or DA |  | n-4 | Drugs: NO6DA, N06DX01 | n-1 |
|  |  |  |  |  | DAS |  | n |  |  |

*LTD: long-term disease status; ICD-10: International Statistical Classification of Diseases and Related Health Problems 10th Revision; GHM: financial hospitalization packages (Groupes Homogène de Malades) ; CCAM: Common classification of medical procedures; LPP: list of reimbursed products; TNB: national table of biology; ATC: Anatomical Therapeutic Chemical ; PD: principal diagnosis; RD: related diagnosis; SAD: associated diagnosis; RUM: Summary of medical unit*

Table S2: Description of respiratory syncytial virus (RSV) hospitalization by age group

|  |  | **50 to 59 years N=1,453 (%)** | **60 to 64 years N=1,242 (%)** | **65 to 74 years N=3,244 (%)** | **75 and above N=9,570 (%)** | **50 and above (all patients)  N=15,509 (%)** |
| --- | --- | --- | --- | --- | --- | --- |
|  | **Month of index hospitalization** |  |  |  |  |  |
|  | July | 16 (1.1) | 13 (1.0) | 23 (0.7) | 39 (0.4) | 91 (0.6) |
|  | August | <10 | <10 | 13 (0.4) | 34 (0.4) | 57 (0.4) |
|  | September | <10 | <10 | 17 (0.5) | 37 (0.4) | 69 (0.4) |
|  | October | 52 (3.6) | 22 (1.8) | 77 (2.4) | 153 (1.6) | 304 (2.0) |
|  | November | 151 (10.4) | 122 (9.8) | 250 (7.7) | 580 (6.1) | 1,103 (7.1) |
|  | December | 384 (26.4) | 357 (28.7) | 839 (25.9) | 2,177 (22.7) | 3,757 (24.2) |
|  | January | 418 (28.8) | 377 (30.4) | 1,072 (33.0) | 3,342 (34.9) | 5,209 (33.6) |
|  | February | 205 (14.1) | 169 (13.6) | 524 (16.2) | 1,861 (19.4) | 2,759 (17.8) |
|  | March | 123 (8.5) | 95 (7.6) | 260 (8.0) | 898 (9.4) | 1,376 (8.9) |
|  | April | 59 (4.1) | 42 (3.4) | 110 (3.4) | 287 (3.0) | 498 (3.2) |
|  | May | 15 (1.0) | 21 (1.7) | 37 (1.1) | 90 (0.9) | 163 (1.1) |
|  | June | 17 (1.2) | 12 (1.0) | 22 (0.7) | 72 (0.8) | 123 (0.8) |
|  | **Area of index hospitalization** |  |  |  |  |  |
|  | Ile de France | 394 (27.1) | 318 (25.6) | 770 (23.7) | 1,957 (20.5) | 3,439 (22.2) |
|  | Auvergne-Rhône Alpes | 170 (11.7) | 165 (13.3) | 505 (15.6) | 1,657 (17.3) | 2,497 (16.1) |
|  | Nouvelle Aquitaine | 137 (9.4) | 109 (8.8) | 321 (9.9) | 1,080 (11.3) | 1,647 (10.6) |
|  | Provence Alpes Cote d'Azur | 109 (7.5) | 100 (8.1) | 239 (7.4) | 908 (9.5) | 1,356 (8.7) |
|  | Normandie | 115 (7.9) | 98 (7.9) | 250 (7.7) | 841 (8.8) | 1,304 (8.4) |
|  | Bourgogne Franche Comté | 73 (5.0) | 69 (5.6) | 188 (5.8) | 637 (6.7) | 967 (6.2) |
|  | Hauts de France | 102 (7.0) | 65 (5.2) | 205 (6.3) | 448 (4.7) | 820 (5.3) |
|  | Bretagne | 73 (5.0) | 71 (5.7) | 165 (5.1) | 496 (5.2) | 805 (5.2) |
|  | Occitanie | 86 (5.9) | 73 (5.9) | 194 (6.0) | 452 (4.7) | 805 (5.2) |
|  | Grand Est | 85 (5.8) | 69 (5.6) | 163 (5.0) | 482 (5.0) | 799 (5.2) |
|  | Pays de Loire | 53 (3.6) | 48 (3.9) | 117 (3.6) | 293 (3.1) | 511 (3.3) |
|  | Centre-Val de Loire | 31 (2.1) | 33 (2.7) | 70 (2.2) | 196 (2.0) | 330 (2.1) |
|  | Outre-mer | 24 (1.7) | 23 (1.9) | 51 (1.6) | 104 (1.1) | 202 (1.3) |
|  | Corse | <10 | 0 (0.0) | <10 | 17 (0.2) | 24 (0.2) |
|  | Missing | 0 | <10 | 0 | <10 | <10 |
|  | **Length of index hospitalization (in days)** |  |  |  |  |  |
|  | Mean (SD) | 13.5 (16.8) | 14.1 (16.1) | 14.7 (16.7) | 15.5 (14.9) | 15.0 (15.6) |
|  | Median (Q1; Q3) | 9.0 (5.0; 15.0) | 10.0 (6.0; 16.0) | 10.0 (6.0; 17.0) | 11.0 (7.0; 18.0) | 11.0 (7.0; 17.0) |
|  | [Min; Max] | [1;261] | [1;193] | [1;238] | [1;304] | [1;304] |
|  | Missing | <10 | <10 | 19 | 50 | 79 |
|  | **Origin of index hospitalization** |  |  |  |  |  |
|  | Emergency room | 866 (66.0) | 713 (64.2) | 2,064 (71.0) | 6,889 (81.7) | 10,532 (76.5) |
|  | Home | 447 (34.0) | 396 (35.7) | 837 (28.8) | 1,471 (17.4) | 3,151 (22.9) |
|  | Nursing home | 0 (0.0) | <10 | <10 | 76 (0.9) | 83 (0.6) |
|  | Missing | 140 | 132 | 337 | 1134 | 1743 |
|  | **Discharge mode from index hospitalization** |  |  |  |  |  |
|  | Home | 1,213 (83.6) | 1,020 (82.3) | 2,539 (78.4) | 6,411 (67.4) | 11,183 (72.4) |
|  | Inpatient death* | 67 (4.6) | 67 (5.4) | 232 (7.2) | 968 (10.2) | 1,334 (8.6) |
|  | Follow-up and rehabilitation care | 60 (4.1) | 56 (4.5) | 176 (5.4) | 912 (9.6) | 1,204 (7.8) |
|  | Other transfer | 107 (7.4) | 88 (7.1) | 251 (7.8) | 701 (7.4) | 1,147 (7.4) |
|  | Nursing home | <10 | <10 | 26 (0.8) | 487 (5.1) | 520 (3.4) |
|  | Home-based hospitalization | <10 | <10 | 14 (0.4) | 35 (0.4) | 55 (0.4) |
|  | Missing | <10 | <10 | <10 | 56 | 66 |
| **Index hospitalization includes a stay in follow-up and rehabilitation care for RSV, N (%)** | | <10 | <10 | 64 (2.0) | 617 (6.4) | 696 (4.5) |
|  | **At least one stay in ICU, N (%)** | 551 (37.9) | 470 (37.8) | 1,145 (35.3) | 1,771 (18.5) | 3,937 (25.4) |

*Information taken from hospital discharge report, differs slightly from the more precise information in Table 3 from healthcare administration system (*Carte Vitale*). SD – Standard Deviation, ICU – Intensive Care Unit

Table S3: Description of respiratory syncytial virus (RSV) hospitalization by comorbidity and immunocompromised status*

|  | **High risk N=2,467 (%)** | **Medium risk N=11,078 (%)** | **Other risks N=751 (%)** | **No comorbidities  N=1,213 (%)** | **All risk groups N=15,509 (%)** |
| --- | --- | --- | --- | --- | --- |
| **Month of index hospitalization** |  |  |  |  |  |
| July | 28 (1.1) | 54 (0.5) | <10 | <10 | 91 (0.6) |
| August | <10 | 42 (0.4) | <10 | <10 | 57 (0.4) |
| September | 11 (0.4) | 52 (0.5) | <10 | <10 | 69 (0.4) |
| October | 63 (2.6) | 204 (1.8) | 11 (1.5) | 26 (2.1) | 304 (2.0) |
| November | 230 (9.3) | 760 (6.9) | 41 (5.5) | 72 (5.9) | 1103 (7.1) |
| December | 627 (25.4) | 2671 (24.1) | 172 (22.9) | 287 (23.7) | 3757 (24.2) |
| January | 787 (31.9) | 3773 (34.1) | 273 (36.4) | 376 (31.0) | 5209 (33.6) |
| February | 386 (15.6) | 1993 (18.0) | 139 (18.5) | 241 (19.9) | 2759 (17.8) |
| March | 181 (7.3) | 995 (9.0) | 66 (8.8) | 134 (11.0) | 1376 (8.9) |
| April | 85 (3.4) | 340 (3.1) | 29 (3.9) | 44 (3.6) | 498 (3.2) |
| May | 35 (1.4) | 113 (1.0) | <10 | <10 | 163 (1.1) |
| June | 25 (1.0) | 81 (0.7) | <10 | 11 (0.9) | 123 (0.8) |
| **Area of index hospitalization** |  |  |  |  |  |
| Ile de France | 778 (31.5) | 2312 (20.9) | 132 (17.6) | 217 (17.9) | 3439 (22.2) |
| Auvergne-Rhône Alpes | 312 (12.6) | 1811 (16.4) | 138 (18.4) | 236 (19.5) | 2497 (16.1) |
| Nouvelle Aquitaine | 217 (8.8) | 1220 (11.0) | 79 (10.5) | 131 (10.8) | 1647 (10.6) |
| Provence Alpes Cote d'Azur | 191 (7.7) | 973 (8.8) | 86 (11.5) | 106 (8.7) | 1356 (8.7) |
| Normandie | 131 (5.3) | 999 (9.0) | 71 (9.5) | 103 (8.5) | 1304 (8.4) |
| Bourgogne Franche Comté | 165 (6.7) | 669 (6.0) | 56 (7.5) | 77 (6.4) | 967 (6.2) |
| Hauts de France | 113 (4.6) | 618 (5.6) | 44 (5.9) | 45 (3.7) | 820 (5.3) |
| Bretagne | 100 (4.1) | 590 (5.3) | 37 (4.9) | 78 (6.4) | 805 (5.2) |
| Occitanie | 122 (4.9) | 565 (5.1) | 44 (5.9) | 74 (6.1) | 805 (5.2) |
| Grand Est | 154 (6.2) | 554 (5.0) | 30 (4.0) | 61 (5.0) | 799 (5.2) |
| Pays de Loire | 101 (4.1) | 357 (3.2) | 16 (2.1) | 37 (3.1) | 511 (3.3) |
| Centre-Val de Loire | 55 (2.2) | 236 (2.1) | 12 (1.6) | 27 (2.2) | 330 (2.1) |
| Outre-mer | 26 (1.1) | 157 (1.4) | <10 | 15 (1.2) | 202 (1.3) |
| Corse | <10 | 15 (0.1) | <10 | <10 | 24 (0.2) |
| Missing | 0 | <10 | 0 | <10 | <10 |
| **Length of index hospitalization (in days)** |  |  |  |  |  |
| Mean (SD) | 15.6 (17.5) | 15.3 (15.5) | 13.5 (12.2) | 12.0 (14.0) | 15.0 (15.6) |
| Median (Q1 ; Q3) | 10.0 (6.0; 18.0) | 11.0 (7.0; 18.0) | 10.0 (6.0; 17.0) | 8.0 (5.0; 14.0) | 11.0 (7.0; 17.0) |
| Min; Max | [1;238] | [1;304] | [1;110] | [1;251] | [1;304] |
| Missing | 23 | 48 | <10 | <10 | 79 |
| **Origin of index hospitalization** |  |  |  |  |  |
| Emergency room | 1141 (51.7) | 7914 (81.0) | 570 (85.6) | 907 (81.0) | 10532 (76.5) |
| Home | 1066 (48.3) | 1789 (18.3) | 89 (13.4) | 207 (18.5) | 3151 (22.9) |
| Nursing home | <10 | 68 (0.7) | <10 | <10 | 83 (0.6) |
| Missing | 258 | 1307 | 85 | 93 | 1743 |
| **Mode of discharge from index hospitalization** |  |  |  |  |  |
| Home | 1916 (77.8) | 7811 (70.8) | 498 (66.9) | 958 (79.5) | 11183 (72.4) |
| Inpatient death^*^ | 210 (8.5) | 1005 (9.1) | 63 (8.5) | 56 (4.6) | 1334 (8.6) |
| Follow-up and rehabilitation care | 142 (5.8) | 917 (8.3) | 71 (9.5) | 74 (6.1) | 1204 (7.8) |
| Other transfer | 167 (6.8) | 849 (7.7) | 53 (7.1) | 78 (6.5) | 1147 (7.4) |
| Nursing home | 15 (0.6) | 415 (3.8) | 55 (7.4) | 35 (2.9) | 520 (3.4) |
| Home-based hospitalization | 14 (0.6) | 33 (0.3) | <10 | <10 | 55 (0.4) |
| Missing | <10 | 48 | <10 | <10 | 66 |
| **Index hospitalization includes a stay in follow-up and rehabilitation care for RSV, N (%)** | 58 (2.4) | 555 (5.0) | 36 (4.8) | 47 (3.9) | 696 (4.5) |
| **At least one stay in ICU, N (%)** | 688 (27.9) | 3030 (27.4) | 70 (9.3) | 149 (12.3) | 3937 (25.4) |
| *Percentages are based on all patients except those with missing values ^*^Inpatient death as recorded in the hospital dicharge record, which may be less precise than information from the DCIR (carte vitale). High risk = immunocompromised patients with at least one comorbidity; medium risk = non-immunocompromised patients with underlying predispositions; other = with generalised risk factors and no comorbidities = patients with no evidence of the former pathologies. SD – Standard Deviation, ICU – Intensive Care Unit* | | | | | |

Table S4: Number of respiratory syncytial virus (RSV) hospitalizations by principal, related and associated diagnosis codes for RSV and co-infections, by epidemiological year

|  | **2015-2016 N=774 (%)** | **2016-2017 N=1,586 (%)** | **2017-2018 N=2,597 (%)** | **2018-2019 N=4,252 (%)** | **2019-2020 N=3,337 (%)** | **2020-2021  N=603 (%)** | **2021-2022 N=2,360 (%)** | **All years N=15,509 (%)** |
| --- | --- | --- | --- | --- | --- | --- | --- | --- |
| **Primary diagnostic codes of hospitalizations for RSV** |  |  |  |  |  |  |  |  |
| J12.1 - Pneumonia due to RSV | 207 (26.7) | 424 (26.7) | 773 (29.8) | 1388 (32.6) | 1,246 (37.3) | 153 (25.4) | 834 (35.3) | 5,025 (32.4) |
| J20.5 - Acute bronchitis due to RSV | 134 (17.3) | 256 (16.1) | 451 (17.4) | 757 (17.8) | 546 (16.4) | 47 (7.8) | 262 (11.1) | 2,453 (15.8) |
| J21.0 - Acute bronchiolitis due to RSV | 16 (2.1) | 45 (2.8) | 85 (3.3) | 123 (2.9) | 79 (2.4) | 13 (2.2) | 88 (3.7) | 449 (2.9) |
| B97.4 - RSV as the cause of diseases classified in other chapters | 0 (0.0) | 0 (0.0) | 0 (0.0) | 0 (0.0) | 0 (0.0) | 0 (0.0) | 0 (0.0) | 0 (0.0) |
| **Related diagnostic codes of hospitalizations for RSV** |  |  |  |  |  |  |  |  |
| J12.1 - Pneumonia due to RSV | <10 | 14 (0.9) | 32 (1.2) | 44 (1.0) | 36 (1.1) | <10 | 35 (1.5) | 175 (1.1) |
| J20.5 - Acute bronchitis due to RSV | <10 | 11 (0.7) | 15 (0.6) | 18 (0.4) | <10 | <10 | <10 | 61 (0.4) |
| J21.0 - Acute bronchiolitis due to RSV | <10 | <10 | <10 | <10 | <10 | <10 | <10 | 16 (0.1) |
| B97.4 - RSV as the cause of diseases classified in other chapters | 0 (0.0) | 0 (0.0) | 0 (0.0) | 0 (0.0) | 0 (0.0) | 0 (0.0) | 0 (0.0) | 0 (0.0) |
| **Associated diagnostic codes of hospitalizations for RSV** |  |  |  |  |  |  |  |  |
| J12.1 - Pneumonia due to RSV | 230 (29.7) | 425 (26.8) | 658 (25.3) | 1,097 (25.8) | 898 (26.9) | 203 (33.7) | 606 (25.7) | 4,117 (26.5) |
| B97.4 - RSV as the cause of diseases classified in other chapters | 86 (11.1) | 225 (14.2) | 438 (16.9) | 760 (17.9) | 597 (17.9) | 189 (31.3) | 478 (20.3) | 2,773 (17.9) |
| J20.5 - Acute bronchitis due to RSV | 162 (20.9) | 343 (21.6) | 475 (18.3) | 741 (17.4) | 466 (14.0) | 46 (7.6) | 287 (12.2) | 2,520 (16.2) |
| J21.0 - Acute bronchiolitis due to RSV | 31 (4.0) | 57 (3.6) | 80 (3.1) | 84 (2.0) | 80 (2.4) | 17 (2.8) | 109 (4.6) | 458 (3.0) |
| **Co-infections (PD, RD or AD)** |  |  |  |  |  |  |  |  |
| Influenza^a^ | 22 (2.8) | 54 (3.4) | 131 (5.0) | 162 (3.8) | 84 (2.5) | <10 | 36 (1.5) | 494 (3.2) |
| COVID-19^b^ | 0 (0.0) | 0 (0.0) | 0 (0.0) | 0 (0.0) | 68 (2.0) | 151 (25.0) | 119 (5.0) | 338 (2.2) |
| *Percentages are based on all patients except those with missing values.* | | | | | | | | |
| *Patients can have more than one diagnosis code during their index hospitalization so column percentages do not add up to 100%.* | | | | | | | | |
| *^a^ICD-10 codes: J09, J10, J11* | | | | | | | | |
| *^b^ICD-10 codes: U07.10, U07.11, U07.14, U07.15* | | | | | | | | |

Table S5: Rates of rehospitalization, admission to nursing homes and mortality within 30 and 90 days of discharge from respiratory syncytial virus (RSV) hospitalization – patients hospitalized for RSV between 2015 and 2022

|  | **High risk:**  **Immune-compromised   N=2,467 (%)** | **Medium risk:**  **Underlying predisposition N=11,078 (%)** | **Other risks N=751 (%)** | **No comorbidities N=1,213 (%)** | **All risk groups N=15,509 (%)** |
| --- | --- | --- | --- | --- | --- |
| **In-hospital death** | 210 (8.5) | 1,002 (9.0) | 59 (7.9) | 55 (4.5) | 1,326 (8.5) |
| **Patients still alive at hospital discharge** | 2,257 (91.5) | 10,076 (91.0) | 692 (92.1) | 1,158 (95.5) | 14,183 (91.5) |
| Admission to a nursing home at 30 days | <10 | 76 (0.8) | 12 (1.7) | 10 (0.9) | 104 (0.7) |
| Admission to a nursing home at 90 days | 12 (0.5) | 224 (2.2) | 30 (4.3) | 15 (1.3) | 281 (2.0) |
| Overnight re-hospitalization for any cause at 30 days | 629 (27.9) | 1,680 (16.7) | 83 (12.0) | 129 (11.1) | 2,521 (17.8) |
| Overnight re-hospitalization for any cause at 90 days | 1,042 (46.2) | 3,128 (31.0) | 142 (20.5) | 225 (19.4) | 4,537 (32.0) |
| Overnight re-hospitalization for respiratory causes at 30 days^a^ | 113 (5.0) | 508 (5.0) | 17 (2.5) | 22 (1.9) | 660 (4.7) |
| Overnight re-hospitalization for respiratory causes at 90 days^a^ | 231 (10.2) | 962 (9.5) | 28 (4.0) | 37 (3.2) | 1,258 (8.9) |
| Overnight re-hospitalization for cardiorespiratory causes at 30 days^b^ | 176 (7.8) | 868 (8.6) | 27 (3.9) | 35 (3.0) | 1,106 (7.8) |
| Overnight re-hospitalization for cardiorespiratory causes at 90 days^b^ | 362 (16.0) | 1,707 (16.9) | 41 (5.9) | 64 (5.5) | 2,174 (15.3) |
| Mortality rate at 30 days | 98 (4.3) | 388 (3.9) | 41 (5.9) | 15 (1.3) | 542 (3.8) |
| Mortality rate at 90 days | 219 (9.7) | 803 (8.0) | 76 (11.0) | 38 (3.3) | 1,136 (8.0) |
| *Percentages are based on all patients except those with missing values.* | | | | | |
| *^a^ICD-10 codes starting with J* | | | | | |
| *^b^ICD-10 codes starting with I or J* | | | | | |

Table S6: Costs of index hospitalization and global healthcare costs (according to national insurance perspective) during reference periods around respiratory syncytial virus (RSV) hospitalization – patients hospitalized for RSV between 2015 and 2022

|  | **High risk: Immune-compromised N=2,467 (%)** | **Medium risk: Underlying predisposition N=11,078 (%)** | **Other risks N=751 (%)** | **No comorbidities N=1,213 (%)** | **All risk groups N=15,509 (%)** |
| --- | --- | --- | --- | --- | --- |
| **Cost of index hospitalization in euros^a^** |  |  |  |  |  |
| Mean (SD) | 9,618 (15,070) | 6,562 (7,739) | 5,290 (6,789) | 5,087 (7,512) | 6,876 (9,346) |
| Median (Q1; Q3) | 4,854 (3,137; 9,534) | 4,289 (3,133; 6,902) | 3,996 (2,924; 5,522) | 3,151 (2,281; 5,086) | 4,252 (3,077; 7,007) |
| Min;Max | [275;252,786] | [279;188,498] | [382;115,102] | [259;109,318] | [259;252,786] |
| Missing | 39 | 239 | 14 | 48 | 340 |
| **Healthcare costs by period, in euros** | | | | | |
| **Pre-infection: 30 days before RSV infection^b^** |  |  |  |  |  |
| Mean (SD) | 5,959 (10,881) | 1,400 (3,153) | 945 (2,242) | 655 (2,062) | 2,041 (5,426) |
| Median (Q1; Q3) | 3,002 (851; 7,951) | 379 (134; 1,072) | 259 (61; 704) | 107 (25; 392) | 436 (132; 1,490) |
| Min;Max | [0;359,717] | [0;57,398] | [0;21,262] | [0;27,925] | [0;359,717] |
| **Pre-hospitalisation: 30 days prior to RSV hospitalisation** |  |  |  |  |  |
| Mean (SD) | 5,295 (6,381) | 1,578 (3,089) | 1,299 (2,478) | 834 (1,978) | 2,092 (3,981) |
| Median (Q1; Q3) | 3,245 (1,101; 7,069) | 549 (196; 1,486) | 383 (120; 1,101) | 197 (56; 684) | 634 (201; 2,078) |
| Min;Max | [0;61,922] | [0;129,619] | [0;21,365] | [0;27,160] | [0;129,619] |
| **Post-hospitalisation: 30 days following RSV hospitalisation^a^** |  |  |  |  |  |
| Mean (SD) | 7,832 (11,510) | 3,757 (8,316) | 3,182 (7,256) | 3,245 (11,204) | 4,336 (9,278) |
| Median (Q1; Q3) | 4,322 (1,503; 9,832) | 985 (385; 3,840) | 644 (244; 2,452) | 416 (154; 1,623) | 1,117 (389; 4,700) |
| Min;Max | [0;151,688] | [0;192,345] | [0;74,326] | [0;192,774] | [0;192,774] |
| **Additional healthcare costs post RSV hospitalisation^a,c^** |  |  |  |  |  |
| Mean (SD) | 1,993 (13,879) | 2,411 (8,377) | 2,242 (7,284) | 2,622 (10,704) | 2,357 (9,648) |
| Median (Q1; Q3) | 445 (-921; 3,729) | 348 (18; 2,146) | 226 (29; 1,230) | 195 (27; 892) | 323 (0; 2292) |
| Min;Max | [-357,893;151,325] | [-43,349;192,345] | [-20,841;73,713] | [-26,460;164,849] | [-357,893;192,345] |
| *Percentages are based on all patients except those with missing values.* *aData for all patients alive at hospital discharge; bPre-infection period: Before RSV infection – From two months before to one month before hospitalisation [-60 days; - 30 days ]. c Additional healthcare costs are the difference between the post-RSV hospitalisation 30-day costs and the 30-day pre-hospitalisation costs and thus could be less than zero. These costs are considered attributable largely to RSV. N are given in column headings, unless otherwise specified. SD – standard deviation, Q1; Q3 – interquartile range.* | | | | | |

Table S7: Model estimates and ANOVA for linear mixed model describing the additional healthcare cost attributable to respiratory syncytial virus (RSV)

|  | **Parameter estimates** | | | | | | **Type 3 ANOVA** |
| --- | --- | --- | --- | --- | --- | --- | --- |
|  | **Categories** | **Estimate (euros)** | **Standard error** | **Lower bound (95%)** | **Upper bound (95%)** | **P(>T)** | **P(>F)** |
| **Reference patient* (intercept)** |  | 2,787 | 465 | 1,876 | 3,698 | <0.0001 |  |
| **Epidemiological year** | 2018-2019 | 0 |  |  |  |  | 0.068 |
|  | 2015-2016 | -86 | 375 | -822 | 649 | 0.818 |  |
|  | 2016-2017 | 452 | 282 | -100 | 1,004 | 0.108 |  |
|  | 2017-2018 | 126 | 234 | -332 | 585 | 0.589 |  |
|  | 2019-2020 | -369 | 217 | -794 | 57 | 0.089 |  |
|  | 2020-2021 | -84 | 419 | -906 | 737 | 0.84 |  |
|  | 2021-2022 | -376 | 242 | -849 | 98 | 0.12 |  |
| **Age category (years)** | 50-59 | 0 |  |  |  |  | 0.392 |
|  | 60-64 | 403 | 383 | -348 | 1,153 | 0.293 |  |
|  | 65-74 | -132 | 310 | -741 | 476 | 0.67 |  |
|  | 75+ | 104 | 280 | -444 | 652 | 0.71 |  |
| **Sex** | Male | 0 |  |  |  |  | 0.015 |
|  | Female | -375 | 154 | -678 | -73 | 0.015 |  |
| **Comorbidity risk group** | No comorbidities | 0 |  |  |  |  | 0.152 |
|  | High risk | -916 | 424 | -1,748 | -84 | 0.031 |  |
|  | Medium risk | -294 | 311 | -904 | 317 | 0.346 |  |
|  | Other risks | -433 | 429 | -1,274 | 408 | 0.313 |  |

* Male patient of 50-59 years without comorbidities, hospitalised for RSV during the 2019-2020 season

Table S8: Inpatient and outpatient costs during reference periods – patients hospitalized for respiratory syncytial virus (RSV) between 2015 and 2022

|  | **50 to 59 years** | | **60 to 64 years** | **65 to 74 years** | **75 and above** | **50 and above** |
| --- | --- | --- | --- | --- | --- | --- |
|  | **N=1,453 (%)** | | **N=1,242 (%)** | **N=3,244 (%)** | **N=9,570 (%)** | **N=15,509 (%)** |
| **INPATIENT HEALTHCARE COSTS, in euros, N^a^** | 1,387 | | 1,174 | 3,014 | 8,608 | 14,183 |
| **Pre-infection: 30 days before RSV infection^b^** |  | |  |  |  |  |
| Mean (SD) | 1,604 (4,906) | | 1,811 (6,756) | 1,584 (7,776) | 915 (3,162) | 1,191 (4,984) |
| Median (Q1; Q3) | 0 (0; 255) | | 0 (0; 579) | 0 (0; 93) | 0 (0; 0) | 0 (0; 0) |
| **Pre-hospitalisation: 30 days prior to RSV hospitalisation^a^** |  | |  |  |  |  |
| Mean (SD) | 1,608 (4,090) | | 1,666 (4,307) | 1,372 (4,089) | 989 (2,758) | 1,182 (3,363) |
| Median (Q1; Q3) | 0 (0; 1,127) | | 0 (0; 1,230) | 0 (0; 801) | 0 (0; 396) | 0 (0; 587) |
| **Post-hospitalisation: 30 days following RSV hospitalisation^a^** |  | |  |  |  |  |
| Mean (SD) | 3,394 (9,867) | | 3,721 (11,484) | 3,207 (8,505) | 3,085 (8,707) | 3,194 (9,046) |
| Median (Q1; Q3) | 0 (0; 2,461) | | 0 (0; 3,278) | 0 (0; 3,143) | 0 (0; 2,986) | 0 (0; 3,021) |
| **Additional healthcare costs post RSV hospitalisation^a,c^** |  | |  |  |  |  |
| Mean (SD) | 1,870 (9,825) | | 1,994 (11,391) | 1,703 (10,981) | 2,223 (8,769) | 2,059 (9,618) |
| Median (Q1; Q3) | 0 (0; 727) | | 0 (0; 1248) | 0 (0; 1,093) | 0 (0; 1,348) | 0 (0; 1,171) |
| **OUTPATIENT HEALTHCARE COSTS, in euros^a^** | | 1,387 | 1,174 | 3,014 | 8,608 | 14,183 |
| **Pre-infection: 30 days before RSV infection^b^** |  | |  |  |  |  |
| Mean (SD) | 1,201 (2,060) | | 1,293 (1,977) | 1,024 (2,021) | 632 (1,187) | 820 (1,580) |
| Median (Q1; Q3) | 464 (71; 1,479) | | 528 (111; 1,595) | 370 (122; 1,003) | 315 (120; 724) | 340 (116; 875) |
| **Pre-hospitalisation: 30 days prior to RSV hospitalisation^a^** |  | |  |  |  |  |
| Mean (SD) | 1,316 (2,078) | | 1,338 (2,048) | 1,054 (2,038) | 674 (1,057) | 867 (1,538) |
| Median (Q1; Q3) | 603 (115; 1,600) | | 604 (161; 1,608) | 440 (159; 1,054) | 370 (156; 794) | 403 (155; 952) |
| **Post-hospitalisation: 30 days following RSV hospitalisation, N^a^** |  | |  |  |  |  |
| Mean (SD) | 1,749 (2,535) | | 1,759 (2,532) | 1,305 (2,315) | 829 (1,218) | 1,097 (1,824) |
| Median (Q1; Q3) | 1,048 (301; 2,209) | | 914 (328; 2,144) | 625 (252; 1,414) | 473 (215; 982) | 554 (233; 1,236) |
| **Additional healthcare costs post RSV hospitalisation, N^a,c^** |  | |  |  |  |  |
| Mean (SD) | 587 (2,162) | | 486 (2,229) | 286 (1,831) | 205 (1,154) | 283 (1,551) |
| Median (Q1; Q3) | 233 (0; 898) | | 202 (-43; 760) | 161 (-73; 576) | 136 (-64; 428) | 151 (-58; 506) |

Percentages are based on all patients except those with missing values. ^a^Data for all patients alive at hospital discharge; ^b^Pre-infection period: Before RSV infection – From two months before to one month before hospitalisation [-60 days; - 30 days ]. c Additional healthcare costs are the difference between the post-RSV hospitalisation 30-day costs and the 30-day pre-hospitalisation costs and thus could be less than zero. These costs are considered attributable largely to RSV. N are given in column headings, unless otherwise specified. SD – standard deviation, Q1; Q3 – interquartile range.
